# The emergence of SARS-CoV-2 variant(s) and its impact on the prevalence of COVID-19 cases in Nabatieh region, Lebanon

**DOI:** 10.1101/2021.04.08.21255005

**Authors:** Fatima Y. Noureddine, Mohamed Chakkour, Ali El Roz, Jana Reda, Reem Al Sahily, Ali Assi, Mohamed Joma, Hassan Salami, Sadek J. Hashem, Batoul Harb, Ali Salami, Ghassan Ghssein

## Abstract

**Background:** An outbreak of an unknown respiratory illness caused by a novel coronavirus, SARS-CoV-2, emerged in the city of Wuhan in Hubei province, China, in December 2019 and was referred to as coronavirus disease-2019 (COVID-19). Soon after, it was declared as a global pandemic by the World Health Organization (WHO) in March 2020. SARS-CoV-2 mainly infects the respiratory tract with different outcomes ranging from asymptomatic infection to severe critical illness leading to death. Different SARS-CoV-2 variants are emerging of which three have raised concerns worldwide due to their high transmissibility among populations.

**Objective:** To study the prevalence of COVID-19 in the region of Nabatieh - South Lebanon during the past year and assess the presence of SARS-CoV-2 variants and their effect on the spread of infection during times of lock-down. Methods: In our study, 37,474 nasopharyngeal swab samples were collected and analyzed for the detection of SARS-CoV-2 virus in suspected patients attending a tertiary health care center in South Lebanon during the period between March 16, 2020 and February 21, 2021.

**Results:** Results demonstrated a variation in the prevalence rates ranging from less than 1% during full lockdown of the country to 8.4% upon easing lockdown restrictions and reaching 27.5% after the holidays and 2021 New Year celebrations. Interestingly, a new variant(s) appeared starting January 2021 with a significant positive association between the prevalence of positive tests and the percentage of the variant(s).

**Conclusion:** Our results indicate that the lockdown implemented by the Lebanese officials presented an effective intervention to contain COVID-19 spread. Our study also showed that lifting lockdown measures during the holidays, which allowed indoor crowded gatherings to occur, caused a surge in COVID-19 cases and rise in the mortality rates nationwide. More importantly, we confirmed the presence of a highly transmissible SARS-CoV-2 variant(s) circulating in the Lebanese community, at least since January 2021 onwards.

## 1. Introduction

In December 2019, several unidentified pneumonia cases surfaced in Wuhan, China, and were linked to a novel coronavirus (CoV) named 2019-nCoV [1], and later, severe acute respiratory syndrome coronavirus 2 (SARS-CoV-2) by the International Committee on Taxonomy of Viruses (ICTV) [2]. The respiratory illness caused by this novel coronavirus was referred to as coronavirus disease-2019 (COVID-19) [3]. COVID-19 was declared a global pandemic and health problem by the World Health Organization (WHO) in March 2020 [4].

Genome-wide phylogenetic analysis of SARS-CoV-2 showed 79.5% sequence identity similarity to severe acute respiratory syndrome coronavirus (SARS-CoV), placing it in the subgenus Sarbecovirus of the genus Betacoronavirus, subfamily Orthocoronavirinae, family Coronaviridae [5, 6]. Coronaviruses are enveloped viruses with positive sense, single-stranded, non-segmented RNA genomes which are, on average, 30 kilobases long [2, 7]. SARS-CoV-2 mainly infects the respiratory tract with different outcomes ranging from asymptomatic infections (1.2%), mild to medium cases (80.9%), severe cases (13.8%), to critical illness (4.7%) and death (2.3%) [8]. According to the daily report of the WHO, as of April 03, 2021, SARS-CoV-2 had affected over 129,619,536 people, and killed more than 2,827,610 people all over the world [9]. Also, a total of 547,727,346 vaccine doses have been administered as of March 30, 2021 [9].

The surveillance of the current infection and the implementation of appropriate medical and governmental policies require early and proper diagnosis of the disease. Molecular detection of SARS-CoV-2 nucleic acid in respiratory samples by one step real-time reverse transcriptase-polymerase chain reaction (RT-PCR) is the gold standard for COVID-19 diagnosis [10]. Most commercially available viral nucleic acid detection kits mainly target three conserved regions of the SARS-CoV-2 genome: the RNA-dependent RNA polymerase (*RdRp*) gene located in the first open reading frame *ORF1ab*; the envelope (*E*) gene; and at a lower rate, the nucleocapsid (*N*) gene, in addition to the spike glycoprotein (*S*)-encoding gene [11]. S glycoprotein is composed of two subunits; a receptor-binding subunit S1 and a membrane-fusion subunit S2, both of which allow SARS-CoV-2 to enter the host cells through utilizing membrane-expressed angiotensin-converting enzyme 2 (ACE-2) receptors [12]. Nonetheless, viruses mutate, and with SARS-CoV-2, different variants have emerged by the end of 2020, of which three have raised concerns worldwide due to their high transmissibility among populations: B.1.1.7, B.1.351, and B.1.1.28 variants. The accumulation of mutations in the *S* gene specifically could alter the viral pathogenicity and immunogenicity. In fact, the emerging SARS-CoV-2 B.1.1.7 variant, carrying the N501Y mutation that alters the conformation of receptor-binding domain (RBD) of the S protein, quickly became the dominant circulating SARS-CoV-2 variant in the United Kingdom in September 2020 [13]. To date, the B.1.1.7 variant has been detected in over 30 countries [13]. S protein mutations are also present in the highly transmissible South African variant (B.1.351) with the 501Y.V2 mutation, and the B.1.1.28 variant (P.1) initially identified in travellers returning from Brazil [14, 15]. Fortunately, experts are highly confident that the efficacy of the available COVID-19 vaccines, especially mRNA-based vaccines, will not be majorly affected by the SARS-CoV-2 variants of concern [16-18].

The first case of COVID-19 was documented in Lebanon on February 21st, 2020 [19]. As of April 03, 2021, 474,925 laboratory confirmed cases including 6,346 deaths had been reported, indicating a rapid spread of the disease across the country [9]. Ten months after the first case was reported, particularly on December 21st, 2020, Lebanon reported the first case of B.1.1.7 SARS-CoV-2 variant infection with growing concerns about the rapid spread of emergent strains and the associated public health implications [20]. Our current study is the first of a kind in Lebanon, since no epidemiological studies have been published, to date, regarding COVID-19 prevalence in Lebanon. The objective of the present study is to assess the prevalence of SARS-CoV-2 and its variant(s) in a Southern Lebanese population and examine whether the new variants are associated with the recent increase in COVID-19 infection rates.

## 2. Materials and Methods

### 2.1 Study design and setting

In this retrospective cohort study, during the period between March 16, 2020 and February 21, 2021, a total of 37,474 nasopharyngeal swab samples were collected and analyzed, by molecular tools, to detect SARS-CoV-2 in suspected patients attending the tertiary health care center “Sheikh Ragheb Harb University Hospital (SRHUH)” in Nabatieh, South Lebanon.

### 2.2 Sample collection and transportation

Swab samples were collected for the extraction of SARS-CoV-2 genome from patients suspected of having COVID-19. Each collected sample was placed in a specialized transport tube containing a sterile solution of normal saline, then transported to the molecular genetics unit at the laboratory of the health care center.

### 2.3 RNA extraction and SARS-CoV-2 detection by qRT-PCR

Upon reception of the samples, RNA extraction was performed either manually using different spin column viral RNA extraction kits, or automatically using the KingFihser™ Flex Purification System (Thermo Scientific™, Thermo Fisher Scientific, USA) with different magnetic beads viral RNA extraction kits (Table S1) according to the manufacturers’ instructions. One-Step Reverse Transcription Real-Time polymerase chain reaction (RT-PCR) was used to confirm the presence of the SARS-CoV-2 RNA in the samples, by amplifying different genes specific for SARS-CoV-2. Different COVID-19 diagnostic kits were used (Table S2). RT-PCR assays were performed through two thermocycler devices: QuantStudio ™ 5 Real-Time PCR System for Human Identification (Applied Biosystem-sTM, Thermo Fisher Scientific, USA) and the Rotor-Gene Q Real-Time PCR Cycler (Qiagen, Germany). The reaction mix contained COVID-19 Reaction Mixture, COVID-19 Probe Mixture, and the RNA sample. Briefly, the following steps were followed in the RT-PCR assays: reverse transcription, 40-45 cycles of denaturation, annealing, extending, and collecting fluorescence signal on different channels. The results were analyzed according to the manufacturers’ instructions.

### 2.4 Prevalence of S-mutant variant(s)

Out of the total number of tests, 6,353 tests were performed using the TaqPath™ COVID-19 CE-IVD RT-PCR Kit (Applied Biosystems™, Thermo Fisher Scientific, USA), that detects the presence of three SARS-CoV-2 genes (*ORF1ab, N* and *S*). The results interpretation was performed according to the provided instructions (Table S3). In case the three genes were detected, SARS-CoV-2 infection by the classic virus was confirmed. However, the detection of two out of three genes, namely the *ORF1ab* and *N* genes, and failure to detect the *S* gene was considered as a finding of possible SARS-CoV-2 mutant strain(s), herein called “*S*-mutant variant(s)”.

## 3. Statistical Analysis

Descriptive statistics were carried out and reported as frequencies and percentages for categorical variables. Quantitative variables were tested for normality distribution using the Kolmogorov–Smirnov test. When data was not normally distributed, Spearman test was used to study the association between the prevalence of positive tests and the percentage of SARS-CoV-2 *S*-mutant variants. If not, Pearson test was conducted. All analyses were performed using SPSS (IBM Corp. Released 2019, SPSS Statistics for Windows Version 26.0, Armonk, NY), and the plots were generated using Origin software (OriginPro, Version 2019b. OriginLab Corporation, Northampton, MA, USA). The level of significance was set at P < 0.05 for all statistical analyses.

## 4. Results

### 4.1 SARS-CoV-2 weekly detection by RT-PCR

Over the period of the study, a total of 37,474 RT-PCR detection tests for SARS-CoV-2 were performed. As shown in Figure 1, COVID-19 testing could be divided into three different stages: the first stage (Stage I) extends between March and the week of July 06th, 2020 where less than 160 tests had been performed weekly; the second stage (Stage II) extends between the week of July 13th and mid-December 2020 revealing an increase in the number of weekly performed tests reaching a maximum of 1,238 tests per week; and the third stage (Stage III) starts in the week of December 21st, 2020 onwards, and it showed a robust increase in the weekly tests performed (up to 2,047 tests per week).

**Figure 1:**
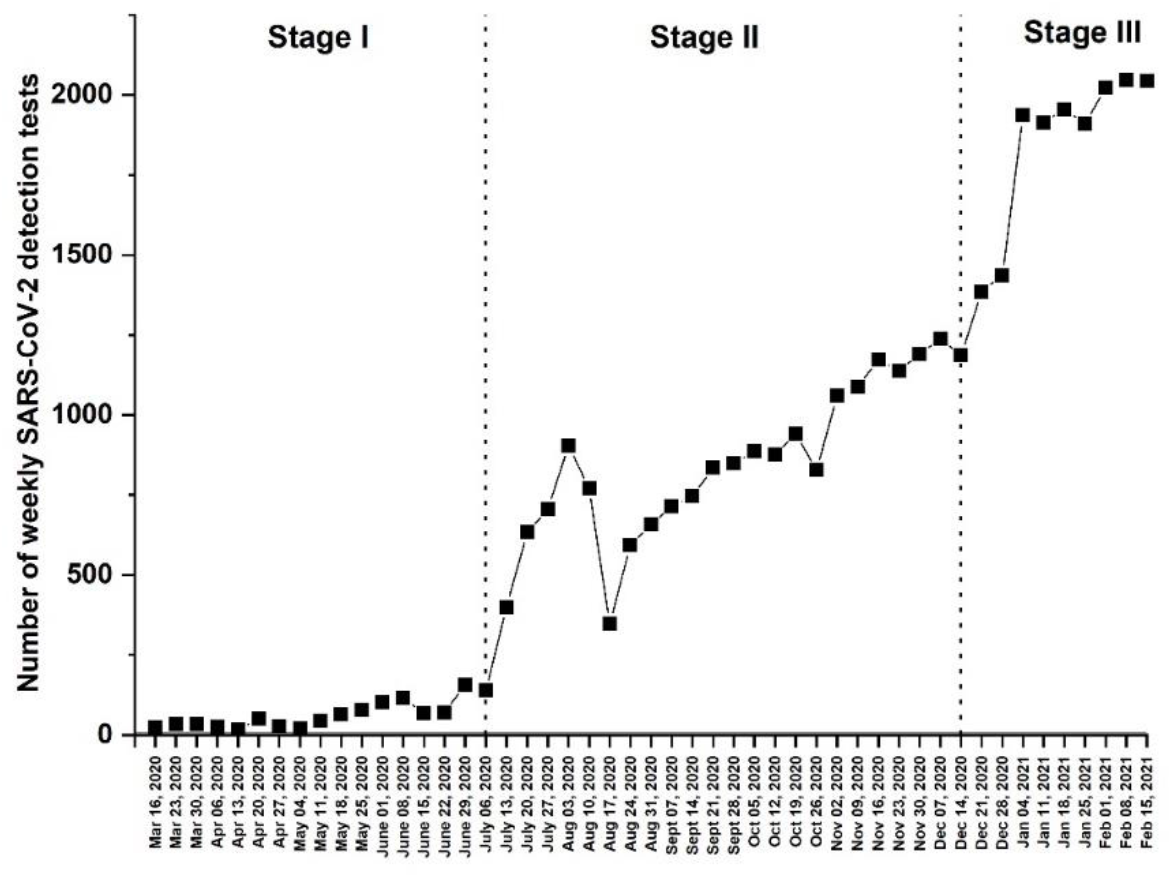
Number of weekly SARS-CoV-2 detection tests performed between March 16, 2020 and February 21, 2021.

### 4.2 Weekly prevalence of SARS-CoV-2 infection

During the period of the study, a total of 6,242 samples (16.6%) rendered positive results for SARS-CoV-2 by RT-PCR, which presents the overall prevalence. However, our results demonstrated a variation in the prevalence rates during the 3 stages previously mentioned (Figure 2). The first stage (Stage I) demonstrated a prevalence rate less than 1% (2/1065), with a weekly percentage fluctuating between 0% and 2.33%. The second stage (Stage II) showed an increase in the prevalence of SARS-CoV-2 positive cases reaching 8.4% (1,670/19,758) with a weekly variation between 0.6% and 16%. In the third stage (Stage III), high prevalence of SARS-CoV-2 positive cases was detected equals to 27.5% (4,570/16,651) with a weekly variation between 19.5% and 33.2% (Figure 2).

**Figure 2:**
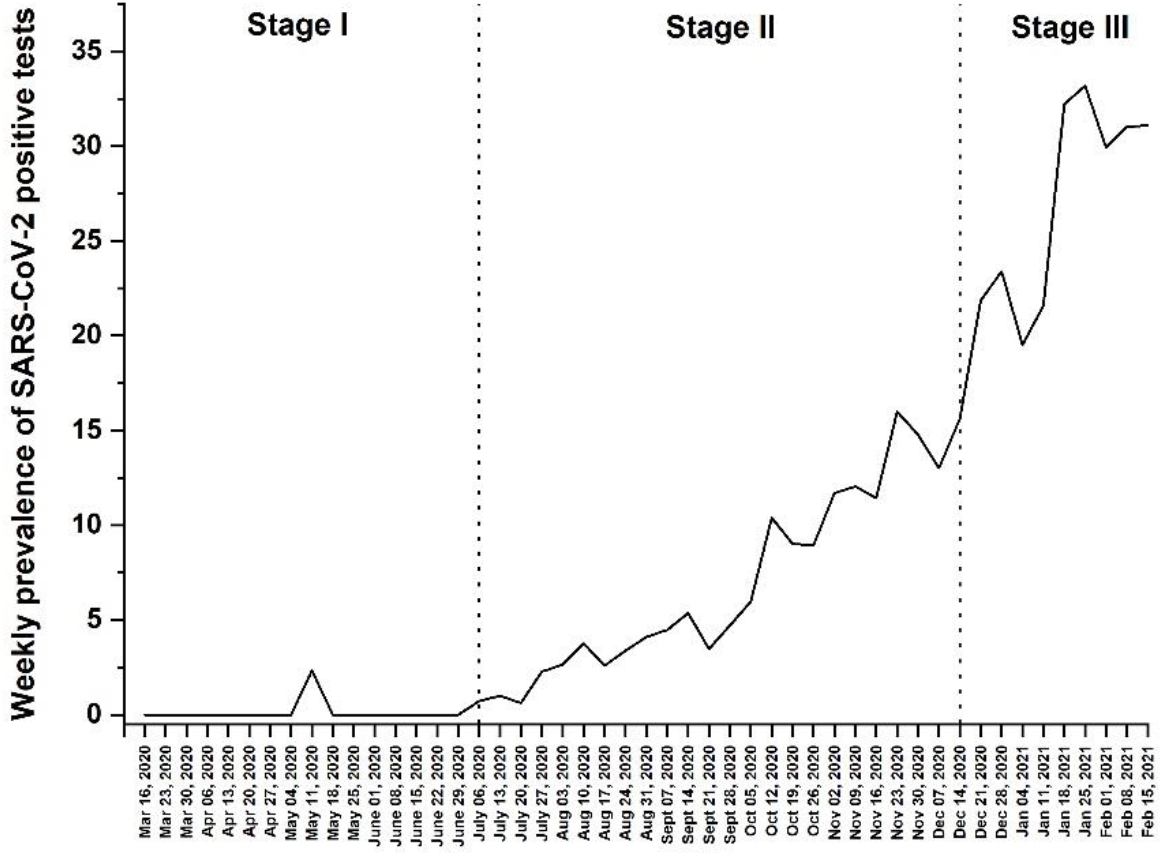
Weekly prevalence of SARS-CoV-2 positive cases between March 16, 2020 and February 21, 2021.

### 4.3 Percentage of SARS-CoV-2 S-mutant variant(s)

Out of the total number of tests performed, 6,353 nasopharyngeal samples were analyzed using the TaqPath COVID-19 CE-IVD RT-PCR Kit that detects the presence of three SARS-CoV-2 genes. Results showed that all positive samples detected in 2020 presented the 3 genes: *ORF1ab, N* and *S* (Figure 3). Starting January 2021, an increase in the percentage of mutant variant(s) (variant(s) lacking the *S* gene prime detected by the kit), herein called “the *S*-mutant variant(s)”, was observed reaching 96.5% in the week of February 15th, 2021 (Figure 3).

**Figure 3:**
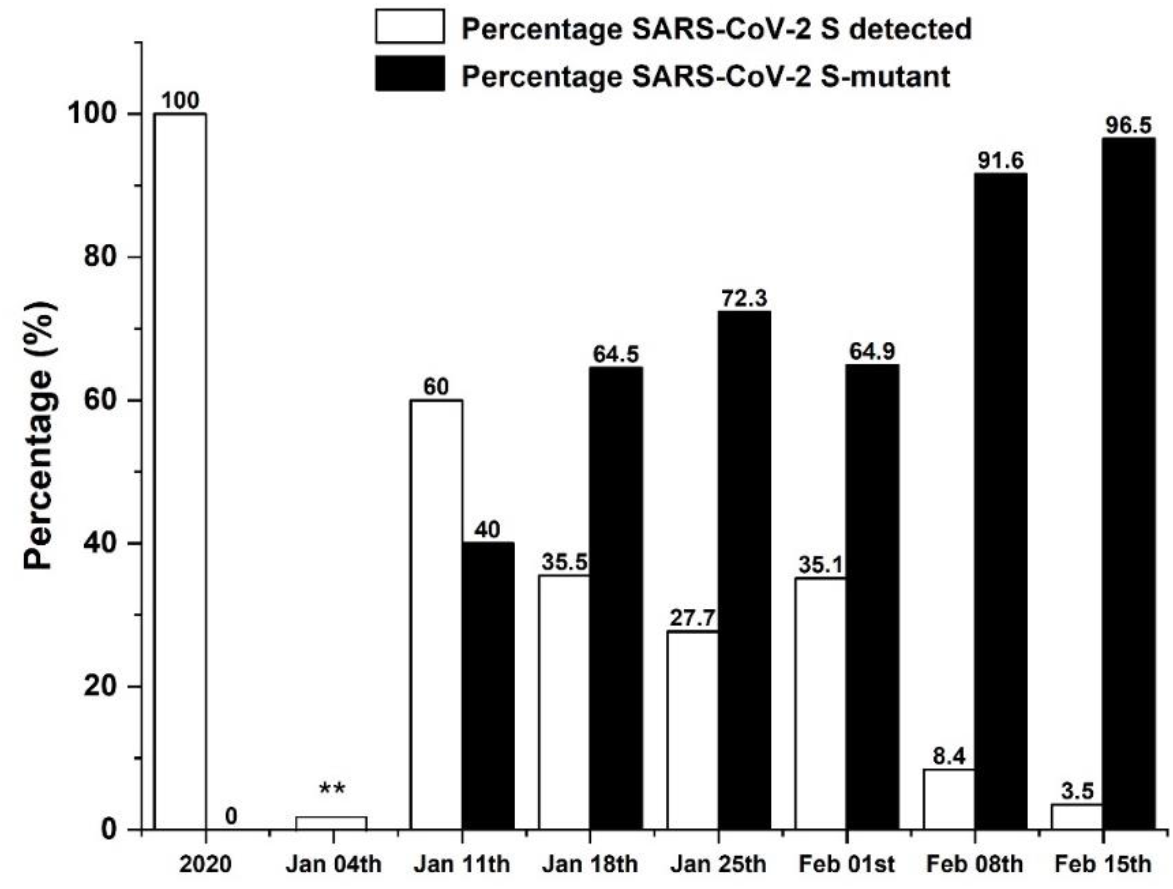
Percentage of classic strain vs. S-mutant variant(s) among SARS-CoV-2 positive cases. **Missing data.

### 4.4 Impact of the SARS-CoV-2 S-mutant strain(s) on the percentage of COVID-19 positive cases

Figure 4 shows the weekly prevalence of COVID-19 positive cases among the 37,474 samples and the weekly percentage of SARS-CoV-2 mutant variants among the samples diagnosed by the TaqPath COVID-19 CE-IVD RT-PCR Kit (6,353 samples). These results showed a remarkable increase in the prevalence of positive cases once the mutant strain(s) were detected starting January 2021.

**Figure 4:**
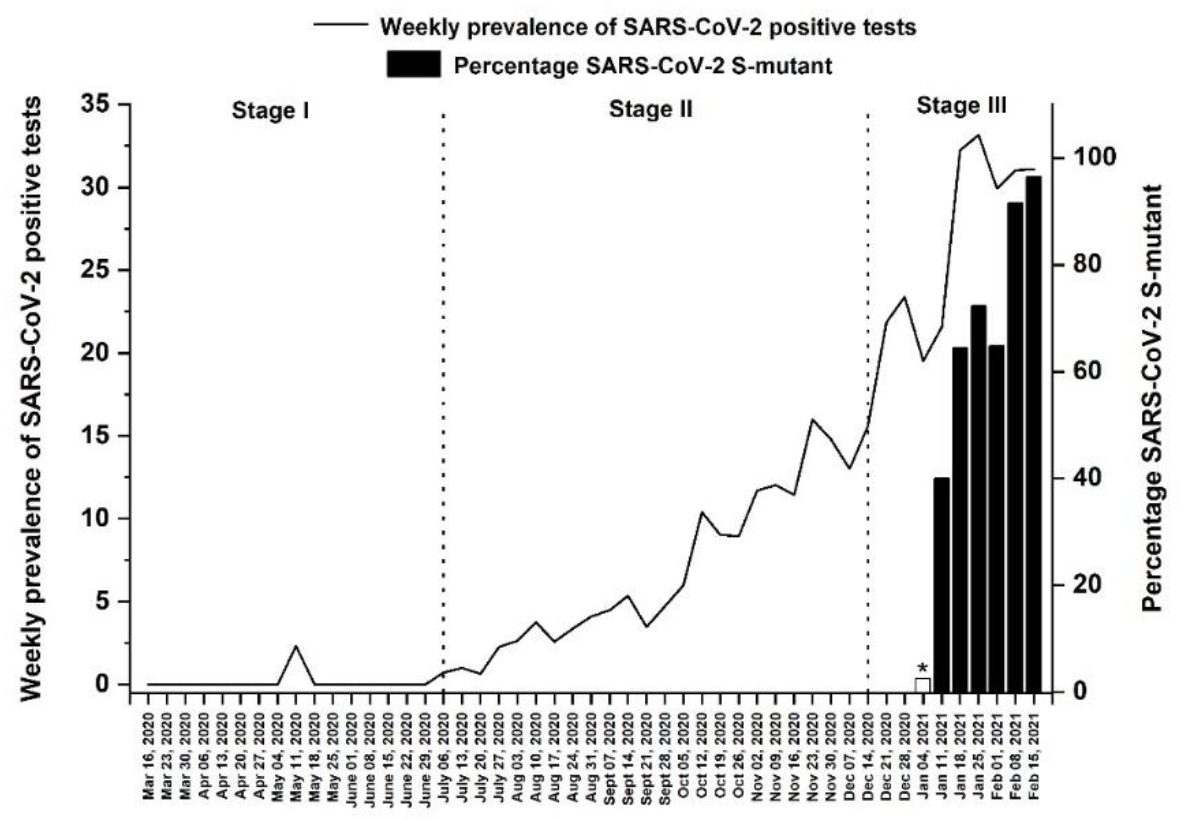
Weekly prevalence of SARS-CoV-2 positive tests and the percentage of S-mutant variant(s) among SARS-CoV-2 positive cases between March 16, 2020 and February 21, 2021. *Missing data.

Using Spearman’s rank-order correlation, we noticed a significant positive association between the prevalence of positive tests and the percentage of *S*-mutants for the period between March 16, 2020 and February 21, 2021 (r = 0.573, P-value < 0.001). Moreover, using Pearson test, a significant positive correlation was also detected in stage III (r = 0.828, P = 0.006).

## 5. Discussion

On February 21st, 2020, the first case of COVID-19 was reported by the Lebanese Ministry of Public Health (MoPH). Few weeks later, the Lebanese officials announced a full lock-down in an early response to control the pandemic where putting the whole country, county or city into full lockdown had been proven elsewhere to be effective in reducing the number of new expected cases [21]. The effectiveness of lockdowns in response to COVID-19 spread was also confirmed in different countries worldwide such as in Italy where the lockdown implemented in March 2020 had a positive impact on containing the spread of the virus [22]. In order to assess the prevalence of COVID-19 positive cases during the whole period (March 2020 – February 2021), our study was divided into 3 stages based on the changes in strictness of measures implemented by health officials to combat the outbreak which fluctuated from extreme stringent to complete easiness during the period of the study; full strict lockdown (Stage I), lifting the lockdown gradually until complete ease of measures (Stage II) and finally the period after holidays where measures are implemented again (Stage III). During stage I of our study during which lockdown was fully implemented (starting March 21, 2020) and before it was partially eased in June 2020, the rate of COVID-19 was very low in South Lebanon, where the number of daily cases ranged between 1 and 10 [23].

Indeed, the results showed a prevalence of COVID-19 positive cases of less than 1% (2/1065) in the studied population during stage I, with a weekly percentage fluctuating between 0% during strict lockdown (March – April) and 2.33% at the end of the first stage where measures started to ease (May – June). This low infection rate can be explained by several reasons including the adherence to the full lockdown measures where people had fear from COVID-19, so they fully abided to the lockdown. In addition, the number of cases in Lebanon during that period was very low [24] which made it easier for health authorities to control the issue. Data from China [25], USA [26] and Indonesia [27] suggest that quarantine and isolation can efficiently reduce the possible peak number of COVID-19 and suspend the time of peak infection. The low positivity rate during stage I of our study could be linked to the low number of tests done during that period (1065) subdivided into less than 160 tests per week. This can be explained by the low number of confirmed COVID-19 cases at the early stages of the outbreak in Southern Lebanon, and by the strict measures taken by municipalities such as early testing for SARS-CoV-2 in individuals with symptoms and early isolation practices which showed to effectively interrupt SARS-CoV-2 transmission [28]. Our results showed less than 1% positivity rate during the strict full lockdown imposed by the health authorities of Lebanon stressing out importance of lockdowns in controlling the spread of the virus. In fact, delaying the lock-down could have resulted in a higher number of cases similar to what happened in other countries where, for instance, delaying lockdown for one week resulted in an 8-32% increase in total COVID-19 cases [29]. Thus, implementing lockdown as early as possible is indispensable to contain the transmission and avoid national outbreaks [21].

The second stage of our study (Stage II) covered the timespan between July and mid-December 2020, a stage during which the Lebanese authorities started easing the lockdown restrictions by the end of June 2020 to reach an almost full back-to-normal-life scenario by December 2020. During this stage, the number of weekly tests done in the health care center included in our present study increased to a maximum of 1,238 tests per week, partnered with a significant increase in the prevalence of positive SARS-CoV-2 cases. In stage II, 1,670 confirmed COVID-19 cases were reported out of 19,758 performed tests with a prevalence rate of SARS-CoV-2 infection of approximately 8.4% with a weekly variation between 0.6% during the first weeks (July 2020) – an expected result since it reveals the outcome of the lockdown – and 16% during the rest of the period (September, October, November, and mid-December 2020) during which restrictions were fully-eased and life started shifting towards normal conditions. The increase in positive cases was expected and it was indeed the result of the ease of the national lockdown measures. In fact, lifting lockdown too early had been shown to cause an increase in infection rates and indeed has led to the second COVID-19 pandemic wave worldwide [30]. For instance, a modelling study performed in France showed that the reduced efficiency of lockdown might be caused by the loose lockdown applied. Dolbeault et al. [31] showed that even when allowing a small margin of the population (2%) to have a higher social interaction level than before lockdown (such as health care workers, supermarket cashiers), the number of infected cases within the population would be higher than in a complete lockdown situation. Rao et al. also demonstrated that if only 50% of the public obeyed lockdown and other precautionary measures implemented by the end of March 2020 in the US, the daily new cases in June would have been 17 times less than the average new COVID-19 cases reported in April (29,000 cases/day). Instead, if a broad, strict lockdown was applied, it could have reduced the daily new cases to 4,300-8,000 cases in May [26]. Moreover, a study performed in England demonstrated that strict lockdown measures imposed by the British authorities on March 23, 2020 led to a significant decrease in COVID-19 cases, hospital admissions, and deaths, while resurrection of COVID-19 cases started to show in the late summer 2020 after most restrictions had been lifted and lockdown was eased [32]. Those studies support our results in which the 8-fold increase in the prevalence of COVID-19 during the second stage (post-lockdown) is highly attributed to the lifting of national lock-down accompanied by an ease in restriction measures. On the other hand, our data shows a peak in the number of tests done during this stage, precisely within the week of August 03, 2020 in which our center witnessed more than 900 tests. In fact, a massive explosion struck in the middle of Beirut within the port, and destroyed most of the city leaving behind more than 200 deaths and more than 7000 injuries [33]. During the days following the explosion, our medical center, similar to most of the Lebanese hospitals, witnessed a huge number of income patients. The center’s strategy during then was to perform PCR test to all admitted patients, a fact which explains the increase in the number of tests during that week.

The third and last stage of our study (stage III) covers the period after mid-December 2020 until 21st of February 2021 (the time of termination of data collection) during which the number of PCR tests performed weekly increased to a maximum of 2,047 tests per week. Out of the 16,651 tests performed during this stage, 4,570 tests were positive, indicating a high prevalence rate of COVID-19 (27.5%) with a weekly variation between ∼20% during December 2020 and approximately 33% during January – mid-February 2021. This increase in the prevalence of SARS-CoV-2 infection during the late weeks of December 2020 and early weeks of January 2021 can be greatly attributed to the holidays’ season during which families and friends gathered in enclosed spaces (houses, restaurants, and even clubs) to celebrate the holidays. Add to that, many clubs and restaurants were fully booked for New Year’s Eve parties and celebrations which made the situation even worse. Data from other countries showed a huge increase in both the prevalence of COVID-19 daily cases and mortality rates after holidays. For example, 200,000 new cases and 3,000 deaths per day were reported in USA alone after thanksgiving celebrations in 2020 [34]. Besides, there was a surge in COVID cases and death rates after 2021 New Year’s gatherings and celebrations [35]. A study performed in 11 European countries suggested an increase in COVID-19 prevalence and mortality rates after holiday vacations during the winter of 2020 [36]. This explains the flood in COVID-19 cases in our study during the early weeks of January as a result of indoor gatherings during the holidays. In fact, viral spread in closed spaces is more likely to occur due to the fact that SARS-CoV-2 is an air-borne virus that is transmitted through air, particularly in crowded and poorly-ventilated places [37].

RNA viruses have an intensely high mutation rate that is correlated with virulence variation and evolution which contributes to viral adaptation [38]. SARS-CoV-2 is an RNA virus, and several testified results indicate that it is rapidly spreading throughout countries and genomes with new mutation hotspots that are emerging, leading to the appearance of new variants that can infect human cells more efficiently, escape neutralizing antibodies and maximize its genome replication [39]. On December 14, 2020, the health authorities in the United Kingdom reported a new SARS-CoV-2 variant namely B.1.1.7, also called as VOC 202012/01 or 201/501 Y.V. This variant was predicted to have emerged somewhere in September 2020 and has become the most dominant SARS-CoV-2 variant circulating in England [13]. Another variant, the 501Y.V2 emerged during the first wave of the South African COVID-19 epidemic in early October 2020, then it spread rapidly to become the dominant lineage in South Africa by the end of November 2020 [40]. A third variant of concern known as the B.1.1.28 variant, also called P.1 variant, was identified in January 2021 and is likely to have originated in Brazil sometime around February 2020 [41]. Nevertheless, in December 21st, 2020, the Lebanese Ministry of Public Health announced the first case of B.1.1.7 SARS-CoV-2 variant infection in a patient arriving from London [20].

On January 9, 2021, the Lebanese authorities re-established a full lockdown across the country in an attempt to contain the outbreak and the hectic increase in infection rates. By the end of January 2021 and beginning of February 2021, our results showed continuous increase in the positivity rates regardless of the lockdown (even 2 weeks after implementing the full lockdown measures). During the first week of February 2021, the prevalence of COVID-19 cases was approximately 32% which is considerably high. Out of the total number of samples, 6,353 were analyzed with the TaqPath COVID-19 CE-IVD RT-PCR Kit which identifies three SARS-CoV-2 genes (*ORFlab, N* and *S*). Since all the previously described variants comprise mutations in the RNA region coding for the viral spike (S) protein [13, 42], we considered the absence of the S-gene in these samples an indicator for the presence of certain SARS-CoV-2 variant or variants. Starting January 2021, the absence of *S* gene in positive samples was detected in our samples. Thus, we postulated the presence of 2 types of variants in our samples; the old one that has been circulating since the beginning of the outbreak in Lebanon, herein referred to as the “classic strain”, and the newly observed variant or variants causing the failure in detecting the *S* gene primer by the kits used in our study, herein referred to as the “*S*-mutant variant(s)”. Our results showed a rapid increase in the prevalence of the “*S*-mutant variant(s)” starting January 2021 where the percentage of its prevalence sharply surged from 72.3% at the end of January 2021, to 96.5% in the week of February 15th, 2021. The prevalence of positive cases during that same period jumped from approximately 20% to 31%. This suggests that the “*S*-mutant variant(s)” variants are indeed highly transmissible and might be responsible for the rapid surge in cases during the end of the third stage in our study. Taking into consideration that all of the aforementioned variants are characterized by being more infectious due to their mutations in the spike (S) protein that gave them an infectious advantage [13, 42], this may explain the rapid increase in COVID-19 positive cases in our samples in correlation with the appearance of the new variants. Also, a significant positive correlation was noted between the prevalence of positive COVID-19 cases and the percentage of “*S*-mutant variant(s)” during Stage III (r = 0. 828, P = 0. 006). Similarly, a study conducted in the UK showed that the SARS-CoV-2 B.1.17 lineage is linked with considerably higher infection and mortality rates among adults [43]. To the best of our knowledge, our results represent the first epidemiological study in Lebanon to demonstrate the presence of new “*S*-mutant variant(s)” within our population, which may be similar to those previously identified in the UK, South Africa and Brazil, or a new one, that is responsible for the surge in COVID-19 cases during the third stage of the study.

Whether the above variant(s) is one of the internationally identified lineages or is a new variant that has emerged within the Lebanese population remains to be deciphered. More studies are needed to investigate the nature of the present variant(s). The lack of appropriate sequencing techniques in our country is a limitation implementing further studies regarding identifying the nature of the new variant(s) due to the financial and economic crisis that the country is currently experiencing. This crisis also prompted the use of different PCR kits to analyse the samples where the market availability of certain kits in Lebanon is indeed challenging. The missing data during the week of January 4^th^, 2021, regarding the percentages of SARS-CoV-2 strains, was due to the unavailability of the TaqPath™ COVID-19 CE-IVD RT-PCR Kit. Finally, a correlation study between demographic and clinical data of the whole 37,474 patients included in the present study will follow in a future study to further explore the effect of SARS-CoV-2 variants in our population.

## 6. Conclusions

Our study emphasizes the importance of strict lockdown measures to control the COVID-19 outbreak and any similar pandemics in the future. It shows how lifting restrictions and allowing indoor gatherings can dramatically influence COVID-19 spread within the population. In addition, our results revealed the presence of novel SARS-CoV-2 variant(s) among the Lebanese population which is likely correlated to the rapid increase in the prevalence rates of COVID-19 positive cases starting of January 2021.

## Data Availability

The data that support the findings of this study are available from the corresponding authors (G.G. and A.S.) upon reasonable request

## Author Contributions

Conceptualization: G.G.; Methodology: F.Y.N., J.R., R.A.S., A.A., M.J., S.J.H. and A.S.; Validation: H.S., A.S. and G.G.; Formal analysis: A.E.R., A.S. and G.G.; Investigation: G.G.; Data curation: F.Y.N., J.R., B.H and A.S.; Software: A.S.; Writing—original draft preparation: F.Y.N., M.C., A.E.R, A.S. and G.G.; Writing—review and editing: All authors; Supervision: H.S., A.E.R, A.S. and G.G.; Project administration: B.H. and H.S.; All authors have read and agreed to the published version of the manuscript.”

## Acknowledgments

We would like to express our gratitude to the healthcare center “Sheikh Ragheb Harb University Hospital” for their support in the conduction of this study.

We thank Dr. Hisham F. Bahmad for taking the time and effort necessary to review the manuscript.

## References

1. Zhu; Zhang, D., Wang, W.; Li, X.; Yang, B.; Song, J.; Zhao, X.; Huang, B.; Shi, W.; Lu, R.; Niu, P.; Zhan, F.; Ma, X.; Wang, D.; Xu, W.; Wu, G.; Gao, G. F.; Tan, W.; A Novel Coronavirus from Patients with Pneumonia in China, 2019. N Engl J Med 2020, 382, (8), 727–733.

2. Gorbalenya, A. E.; Gulyaeva, A. A.; Lauber, C.; Gorbalenya, A. E.; Leontovich, A. M.; Penzar, D.; Samborskiy, D.; Baker, S. C.; Baric, R. S.; de Groot, R. J.; Drosten, C.; Haagmans, B. L.; Neuman, B. W.; Perlman, S.; Poon, L. L. M.; Sola, I.; Ziebuhr, J.; Sidorov, I. A.; Severe acute respiratory syndrome-related coronavirus: The species and its viruses - a statement of the Coronavirus Study Group. bioRxiv 2020.

3. WHO (2019). “Novel-Coronavirus-2019”. https://www.who.int/es/emergencies/diseases/novel-coronavirus-2019.

4. WHO (2020). “WHO Director-General’s opening remarks at the media briefing on COVID-19-11 March 2020”. (Geneva: World Health Organization).

5. Lu, R.; Zhao, X.; Li, J.; Niu, P.; Yang, B.; Wu, H.; Wang, W.; Song, H.; Huang, B.; Zhu, N.; Bi, Y.; Ma, X.; Zhan, F.; Wang, L.; Hu, T.; Zhou, H.; Hu, Z.; Zhou, W.; Zhao, L.; Chen, J.; Meng, Y.; Wang, J.; Lin, Y.; Yuan, J.; Xie, Z.; Ma, J.; Liu, W. J.; Wang, D.; Xu, W.; Holmes, E. C.; Gao, G. F.; Wu, G.; Chen, W.; Shi, W.; Tan, W.; Genomic characterisation and epidemiology of 2019 novel coronavirus: implications for virus origins and receptor binding. The lancet. 2020, 395, (10224), 565–574.

6. Hasoksuz, M.; Kilic, S.; Sarac, F.; Coronaviruses and sars-cov-2. Turk. J. Med. Sci. 2020, 50, (SI-1), 549–556.

7. Jin, Y.; Yang, H.; Ji, W.; Wu, W.; Chen, S.; Zhang, W.; Duan, G.; Virology, Epidemiology, Pathogenesis, and Control of COVID-19. Viruses Viruses 2020, 12, (4), 372.

8. Epidemiology Working Group for Ncip Epidemic Response, C. C. f. D. C.,Prevention, [The epidemiological characteristics of an outbreak of 2019 novel coronavirus diseases (COVID-19) in China]. Zhonghua liuxingbingxue zazhi 2020, 41, (2), 145–151.

9. WHO, “WHO Coronavirus (COVID-19) Dashboard.” https://covid19.who.int/.

10. Böger, B.; Fachi, M. M.; Vilhena, R. O.; Cobre, A. F.; Tonin, F. S.; Pontarolo, R.; Systematic review with meta-analysis of the accuracy of diagnostic tests for COVID-19. American Journal of Infection Control 2021, 49, (1), 21–29.

11. Taleghani, N.; Taghipour, F.; Diagnosis of COVID-19 for controlling the pandemic: A review of the state-of-the-art. Biosens. Bioelectron. 2021, 174.

12. Lokman, S. M.; Rasheduzzaman, M.; Salauddin, A.; Barua, R.; Tanzina, A. Y.; Rumi, M. H.; Hossain, M. I.; Mannan, A.; Hasan, M. M.; Siddiki, A. M. A. M. Z.; Hasan, M. M.; Exploring the genomic and proteomic variations of SARS-CoV-2 spike glycoprotein: A computational biology approach. Infec. Genet. Evol. 2020, 84.

13. Galloway, S. E.; Paul, P.; MacCannell, D. R.; Johansson, M. A.; Brooks, J. T.; MacNeil, A.; Slayton, R. B.; Tong, S.; Silk, B. J.; Armstrong, G. L.; Biggerstaff, M.; Dugan, V. G.; Emergence of SARS-CoV-2 B.1.1.7 Lineage - United States, December 29, 2020-January 12, 2021. Morbidity and mortality weekly report 2021, 70, (3), 95–99.

14. Tang, J. W.; Toovey, O. T. R.; Harvey, K. N.; Hui, D. D. S.; Introduction of the South African SARS-CoV-2 variant 501Y.V2 into the UK. Journal of Infection Journal of Infection 2021.

15. Fujino, T.; Nomoto, H.; Kutsuna, S.; Ujiie, M.; Suzuki, T.; Sato, R.; Fujimoto, T.; Kuroda, M.; Wakita, T.; Ohmagari, N.; Novel SARS-CoV-2 Variant in Travelers from Brazil to Japan. Emerg Infect Dis 2021, 27, (4).

16. Moore, J. P.; Approaches for Optimal Use of Different COVID-19 Vaccines: Issues of Viral Variants and Vaccine Efficacy. JAMA 2021.

17. Shen, X.; Tang, H.; McDanal, C.; Wagh, K.; Fischer, W. M.; Theiler, J.; Yoon, H.; Li, D.; Haynes, B. F.; Saunders, K. O.; Gnanakaran, S.; Hengartner, N. W.; Pajon, R.; Smith, G.; Dubovsky, F.; Glenn, G. M.; Korber, B. T.; Montefiori, D. C.; SARS-CoV-2 Variant B.1.1.7 is Susceptible to Neutralizing Antibodies Elicited by Ancestral Spike Vaccines. SSRN Journal 2021.

18. Madhi, S. A.; Baillie, V.; Cutland, C. L.; Voysey, M.; Koen, A. L.; Fairlie, L.; Padayachee, S. D.; Dheda, K.; Barnabas, S. L.; Bhorat, Q. E.; Briner, C.; Kwatra, G.; Ahmed, K.; Aley, P.; Bhikha, S.; Bhiman, J. N.; Bhorat, A. E.; du Plessis, J.; Esmail, A.; Groenewald, M.; Horne, E.; Hwa, S. H.; Jose, A.; Lambe, T.; Laubscher, M.; Malahleha, M.; Masenya, M.; Masilela, M.; McKenzie, S.; Molapo, K.; Moultrie, A.; Oelofse, S.; Patel, F.; Pillay, S.; Rhead, S.; Rodel, H.; Rossouw, L.; Taoushanis, C.; Tegally, H.; Thombrayil, A.; van Eck, S.; Wibmer, C. K.; Durham, N. M.; Kelly, E. J.; Villafana, T. L.; Gilbert, S.; Pollard, A. J.; de Oliveira, T.; Moore, P. L.; Sigal, A.; Izu, A.; Group, N.-S. G. W.-V. C.; Efficacy of the ChAdOx1 nCoV-19 Covid-19 Vaccine against the B.1.351 Variant. The New England journal of medicine 2021.

19. MOPH, “novel-coronavirus-2019.” https://www.moph.gov.lb/en/Pages/2/24870/novel-coronavirus-2019.

20. “Global Report Investigating Novel Coronavirus Haplotypes.” Accessed: Mar. 09, 2021. https://cov-lineages.org/global_report_B.1.1.7.html.

21. Johanna, N.; Citrawijaya, H.; Wangge, G.; Mass screening vs lockdown vs combination of both to control COVID-19: A systematic review. Journal of public health research 2020, 9, (4).

22. Signorelli, C.; Odone, A.; Signorelli, C.; Scognamiglio, T.; Odone, A.; COVID-19 in Italy: Impact of containment measures and prevalence estimates of infection in the general population. Acta Biomed. 2020, 91, 175–179.

23. PCM, “Disaster Risk Management Unit of Lebanon - COVID-19 Daily Situation Reports,” 2020. http://drm.pcm.gov.lb/.

24. Lesotho: the latest coronavirus counts, charts and maps,” 2021. https://graphics.reuters.com/world-coronavirus-tracker-and-maps/countries-and-territories/lesotho/.

25. Hou, C.; Chen, J.; Zhou, Y.; Hua, L.; Yuan, J.; He, S.; Guo, Y.; Zhang, S.; Jia, Q.; Zhao, C.; Zhang, J.; Xu, G.; Jia, E.; The effectiveness of quarantine of Wuhan city against the Corona Virus Disease 2019 (COVID-19): A well-mixed SEIR model analysis. Journal of medical virology 2020, 92, (7), 841–848.

26. Srinivasa Rao, A.S.R., Krantz, S. G.; Continued and Serious Lockdown Could Minimize Many Newly Transmitted Cases of COVID-19 in the U.S.: Wavelets, Deterministic Models, and Data. medRxiv 2020, 2020.04.30.20080978.

27. Putra, Z. A.; Abidin, S. A. Z.; Application of seir model in covid-19 and the effect of lockdown on reducing the number of active cases. Indones. J. Sci. Technol. 2020, 5, (2), 185–192.

28. Velavan, T. P.; Meyer, C. G.; COVID-19: A PCR-defined pandemic. International Journal of Infectious Diseases 2021, 103, 278–279.

29. Tellis, G. J.; Sood, A.; Sood, N.; Price of Delay in Covid-19 Lockdowns: Delays Spike Total Cases, Natural Experiments Reveal. SSRN Journal 2020.

30. Tang, B.; Xia, F.; Tang, S.; Bragazzi, N. L.; Li, Q.; Sun, X.; Liang, J.; Xiao, Y.; Wu, J.; The effectiveness of quarantine and isolation determine the trend of the COVID-19 epidemics in the final phase of the current outbreak in China. International Journal of Infectious Diseases 2020, 95, 288–293.

31. Dolbeault, J.; Turinici, G.; Social heterogeneity and the COVID-19 lockdown in a multi-group SEIR modeln. medRxiv 2021, 2020.05.15.20103010.

32. Davies, N. G.; Barnard, R. C.; Jarvis, C. I.; Russell, T. W.; Semple, M. G.; Jit, M.; Edmunds, W. J.; Centre for Mathematical Modelling of Infectious Diseases, C.-W. G.,investigators, I. C., Association of tiered restrictions and a second lockdown with COVID-19 deaths and hospital admissions in England: a modelling study. The Lancet. Infectious diseases 2021, 21, (4), 482–492.

33. L. Unicef, “Beirut Explosions | UNICEF Lebanon” 2020. https://www.unicef.org/lebanon/beirut-explosions.

34. Plater, R. The Post-Thanksgiving COVID-19 Surge Is Here: What to Expect Now; 2020.

35. Bryant, M.; US braces for post-holiday Covid surge as death toll passes 350,000. The Guardian 2021.

36. Björk, J.; Mattisson, K.; Ahlbom, A.; Impact of winter holiday and government responses on mortality in Europe during the first wave of the COVID-19 pandemic. European journal of public health 2021.

37. Lewis, D.; COVID-19 rarely spreads through surfaces. So why are we still deep cleaning? Nature 2021, 590, (7844), 26–28.

38. Duffy, S.; Why are RNA virus mutation rates so damn high? PLoS Biology 2018, 16, (8).

39. Pachetti, M.; Storici, P.; Masciovecchio, C.; Pachetti, M.; Giudici, F.; Marini, B.; Mauro, E.; Ippodrino, R.; Benedetti, F.; Zella, D.; Angeletti, S.; Ciccozzi, M.; Gallo, R. C.; Gallo, R. C.; Zella, D.; Emerging SARS-CoV-2 mutation hot spots include a novel RNA-dependent-RNA polymerase variant. J. Transl. Med. 2020, 18, (1).

40. Tegally, H.; Wilkinson, E.; Giovanetti, M.; Iranzadeh, A.; Fonseca, V.; Giandhari, J.; Doolabh, D.; Pillay, S.; San, E. J.; Msomi, N.; Mlisana, K.; von Gottberg, A.; Walaza, S.; Allam, M.; Ismail, A.; Mohale, T.; Glass, A. J.; Engelbrecht, S.; Van Zyl, G.; Preiser, W.; Petruccione, F.; Sigal, A.; Hardie, D.; Marais, G.; Hsiao, M.; Korsman, S.; Davies, M.-A.; Tyers, L.; Mudau, I.; York, D.; Maslo, C.; Goedhals, D.; Abrahams, S.; Laguda-Akingba, O.; Alisoltani-Dehkordi, A.; Godzik, A.; Wibmer, C. K.; Sewell, B. T.; Lourenço, J.; Alcantara, L. C. J.; Pond, S. L. K.; Weaver, S.; Martin, D.; Lessells, R. J.; Bhiman, J. N.; Williamson, C.; de Oliveira, T.; Emergence and rapid spread of a new severe acute respiratory syndrome-related coronavirus 2 (SARS-CoV-2) lineage with multiple spike mutations in South Africa. medRxiv 2020.

41. Candido, D. S.; Claro, I. M.; de Jesus, J. G.; Souza, W. M.; Moreira, F. R. R.; Dellicour, S.; Mellan, T. A.; du Plessis, L.; Pereira, R. H. M.; Sales, F. C. S.; Manuli, E. R.; Thézé, J.; Almeida, L.; Menezes, M. T.; Voloch, C. M.; Fumagalli, M. J.; Coletti, T. s. M.; da Silva, C. A. M.; Ramundo, M. S.; Amorim, M. R.; Hoeltgebaum, H. H.; Mishra, S.; Gill, M. S.; Carvalho, L. M.; Buss, L. F.; Prete, C. A.; Ashworth, J.; Nakaya, H. I.; Peixoto, P. S.; Brady, O. J.; Nicholls, S. M.; Tanuri, A.; Rossi, A. t. D.; Braga, C. K. V.; Gerber, A. L.; de C. Guimarães, A. P.; Gaburo, N.; Alencar, C. S.; Ferreira, A. C. S.; Lima, C. X.; Levi, J. E.; Granato, C.; Ferreira, G. M.; Francisco, R. S.; Granja, F.; Garcia, M. T.; Moretti, M. L.; Perroud, M. W.; Castiñeiras, T. M. P. P.; Lazari, C. S.; Hill, S. C.; de Souza Santos A. A.; Simeoni, C. L.; Forato, J.; Sposito, A. C.; Schreiber, A. Z.; Santos, M. N. N.; de SÁ, C. Z.; Souza, R. P.; Resende-Moreira, L. C.; Teixeira, M. M.; Hubner, J.; Leme, P. A. F.; Moreira, R. G.; Nogueira, M. c. L.; Ferguson, N. M.; Costa, S. F.; Proenca-Modena, J. L.; Vasconcelos, A. T. R.; Bhatt, S.; Lemey, P.; Wu, C.-H.; Rambaut, A.; Loman, N. J.; Aguiar, R. S.; Pybus, O. G.; Sabino, E. C.; Faria, N. R.; Evolution and epidemic spread of SARS-CoV-2 in Brazil. Science Science 2020, 369, (6508), 1255–1260.

42. Nonaka, C. K. V.; Franco, M. M.; Gräf, T.; de Lorenzo Barcia, C. A.; de Ávila Mendonça, R. N.; de Sousa, K. A. F.; Neiva, L. M. C.; Fosenca, V.; Mendes, A. V. A.; de Aguiar, R. S.; Giovanetti, M.; de Freitas Souza B S, Genomic Evidence of SARS-CoV-2 Reinfection Involving E484K Spike Mutation, Brazil. Emerging infectious diseases 2021, 27, (5).

43. Challen, R.; Brooks-Pollock, E.; Read, J. M.; Dyson, L.; Tsaneva-Atanasova, K.; Danon, L.; Risk of mortality in patients infected with SARS-CoV-2 variant of concern 202012/1: matched cohort study. BMJ 2021.

